# Predicting ADC Map Quality from T2-Weighted MRI: A Deep Learning Approach for Early Quality Assessment to Assist Point-of-Care

**DOI:** 10.1101/2025.01.15.25320592

**Authors:** Jeffrey R. Brender, Mitsuki Ota, Nathan Nguyen, Joshua W. Ford, Shun Kishimoto, Stephanie A. Harmon, Bradford J. Wood, Peter A. Pinto, Murali Cherukuri Krishna, Peter L. Choyke, Baris Turkbey

## Abstract

**Purpose:** Poor quality prostate MRI images, especially ADC maps, can lead to missed lesions and unnecessary repeat scans. To address this issue, we aimed to develop an automated method to predict ADC map quality from T2 images acquired earlier in the scanning process.

**Materials and Methods:** A paired multi-site image corpus of T2-weighted images and ADC maps was constructed from 486 patients imaged in-house and at 62 external clinics. A senior radiologist assigned 1-3 quality ratings to each image set, later converted to a binary “non-diagnostic” or “diagnostic” scale. A deep learning model and a rectal cross-sectional area measurement approach were developed to predict ADC image quality from T2 images. Model performance was evaluated retrospectively by accuracy, sensitivity, negative and positive predictive value, and AUC.

**Results:** No single acquisition parameter in the metadata was statistically associated with image quality for either T2 or ADC maps. Quality scores of the same modality showed low correlation across sites (r∼0.2). In the challenging task of predicting ADC quality from prior T2 images, our model achieved performance comparable to current single-site models directly using ADC maps, with 83% sensitivity and 90% negative predictive value. The model showed stronger performance on in-house data (94±2% accuracy) despite being trained exclusively on multicenter external data. Rectal cross-sectional area on T2 images provided an interpretable quality metric (AUC 0.65).

**Conclusion:** The probability of low quality, uninterpretable ADC maps can be inferred early in the imaging process by neural network approach, allowing corrective action to be employed.

## Introduction

Accurate diagnosis of localized prostate cancer relies heavily on high-quality multi-parametric MRI (mpMRI). Because of its high negative predictive value when performed properly and reasonable cancer detection rate, mpMRI is usually the first, crucial step in the diagnostic pathway in screening positive patients (1). Because MRI is commonly used to guide prostate biopsies, poor quality images heavily influence all downstream events. Suboptimal mpMRI image quality can lead to delayed diagnosis, unnecessary biopsies, and misclassification of tumors, ultimately compromising patient care. A 2023 multicenter European study revealed that low-quality images were over three times more likely to be upgraded to a higher-grade, more dangerous status after biopsy (2). 40% of images in this study fell into the low-quality category, indicating a significant number of potentially cancer harboring lesions may remain undetected by mpMRI due to image quality issues (2). In the best case, a clinically nondiagnostic MRI requires another visit for repeat imaging, which is a misallocation of scarce resources. In the worst case, a suboptimal image misses the timely diagnosis of a potentially fatal cancer.

Efforts to enhance prostate image quality can be divided into quality control measures, focusing on establishing processes for high image quality assurance, and quality assessment, aiming to identify low-quality images early in the diagnostic process (1). As a quality control initiative, the PI-RADS committee and the later PI-QUAL standard sought to standardize imaging protocols by setting minimum technical requirements for MRI acquisition, aiming to reduce variability between imaging sites (3, 4). The success of these initiatives has been a subject of debate, with some studies indicating continuous improvement as PI-RADSv2 technical requirements are met, (5) while others suggest a weak correlation between radiologist-assessed image quality and adherence to these standards, either individually or collectively (6). While quality control methods will undoubtedly continue to be essential for meeting quality standards and will likely improve as the standards continue to evolve (7, 8), quality assessment will remain a vital part of the imaging assessment pipeline (9).

Quality control can potentially be assessed manually by the radiologist on duty. However, the normal operating practice of hospitals makes this difficult to achieve in a timely manner. Automatic quality assessment by artificial intelligence can potentially alert the technologist to methodological problems and allow them to address these issues before the patient leaves the imaging facility (10–12). Automatic quality assessment currently suffers from two issues that may limit its clinical effectiveness. Although high accuracy in binary quality classification of T2 and ADC images has been achieved in a single-site setting with consistent protocols using a convolutional neural network (10), replicating this accuracy across multiple sites has proved challenging (12). Second, the ADC map must first be acquired to be assessed, which does not necessarily contribute to point-of-care decision-making and will still require repeat imaging for patients on another day. Within mpMRI, the ADC maps from diffusion weighted imaging (DWI) have been particularly valuable in identifying cancer lesions. However, the EPI sequence used in DWI is susceptible to magnetic susceptibility distortion which can distort the posterior region of the prostate, where majority of clinically significant lesions are expected to be located. Given that acquiring the ADC map is the most time-consuming step of mpMRI and occurs late in the imaging procedure, an interventional method capable of predicting the quality of the ADC map before DWI acquisition occurs would have obvious benefits. In this study, we first demonstrate the limitations of standardizing technical parameters, finding no single acquisition parameter consistently predicts image quality. Given these constraints, we developed a deep learning approach using a multi-site training corpus which evaluates T2 images to predict the future quality of the ADC maps, allowing corrective actions before lengthy DWI sequences.

## Methods

### Study Population

A multi-site training corpus of mpMRI images was retrospectively constructed from 486 patients (mean age 64.0 ± 7.3 SD, mean weight 85.3 kg ± 15.4 SD) imaged first at one of 62 different institutions before being subsequently referred to our facility for prostate imaging (13). The study was conducted under Institutional Review Board approval and in compliance with HIPAA regulations, with all participants providing written informed consent (<u>ClinicalTrials.gov</u> identifier: NCT03354416). In compliance with privacy regulations, all DICOM images underwent a two-step anonymization process: automated removal of header metadata followed by manual verification to ensure the absence of protected health information in both file names and image content. This unique paired dataset encompasses a wide array of imaging procedures, scanning hardware, and pulse sequences, providing a comprehensive reflection of the clinical imaging landscape.

Image quality was assessed by a radiologist (*, with over 15 years of experience in prostate cancer imaging) for both T2 images and ADC maps, who evaluated both technical distortions (e.g., motion, artifacts, noise, and aliasing) and perceptual issues (e.g., blurred prostate capsule or zones, unclear external urethral sphincter, or excessive rectal gas). The used quality evaluation criteria matched PIQUALv2.(14) Each image set received a 1-3 rating: 1 for poor quality (significantly hindering diagnosis), 2 for adequate quality (diagnostically usable despite minor distortions), and 3 for high quality (no distortions) (Fig. 1). To simplify analysis, the “high quality” and “adequate” categories were combined into a single “diagnostic” category for a binary quality classification system.

**Figure 1:**
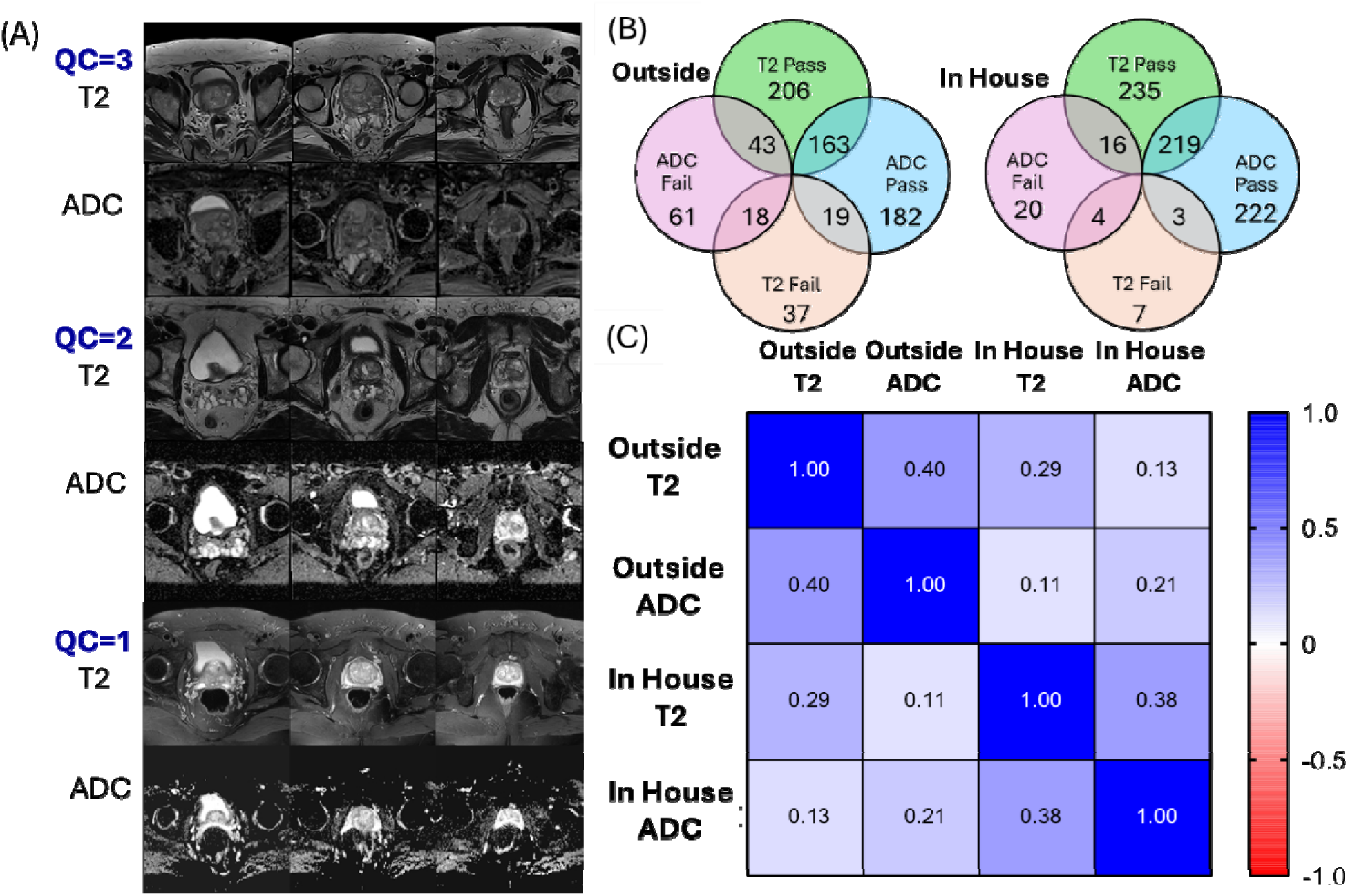
Quality assessment of prostate MRI across institutions. (A) Representative axial T2-weighted images and corresponding ADC maps demonstrating the three-point quality scoring system (QC=3: optimal quality, QC=2: adequate quality, QC=1: non-diagnostic quality). (B) Venn diagrams showing the distribution of image quality between in house and outside institutions for both T2 and ADC sequences. (C) Correlation matrix between T2 and ADC quality scores for the same patient within and across institutions

### Neural Network Building

To develop our predictive model, we implemented a multi-scale deep learning approach analyzing 14 consecutive axial slices centered on the imaging isocenter, which was assumed to be centered on the prostate midgland. Images were divided into three anatomically relevant, overlapping regions (bladder, prostate, and rectum) (Fig. 2B). The validation process employed a rigorous multi-stage approach to ensure unbiased model evaluation (Fig. 2B). First, 20% of the study IDs were randomly held out as a validation set. The remaining 80% underwent stratified 5-fold cross-validation (80/20), with training data for the neural network incorporating both NIH and external images while test data consisted exclusively of external images. A logistic regression model as described below was then trained on the test set predictions using 5-fold cross-validation for parameter optimization. The final performance metrics were obtained by applying this regression model to the completely independent validation set, using a classification threshold of 0.5.

**Figure 2:**
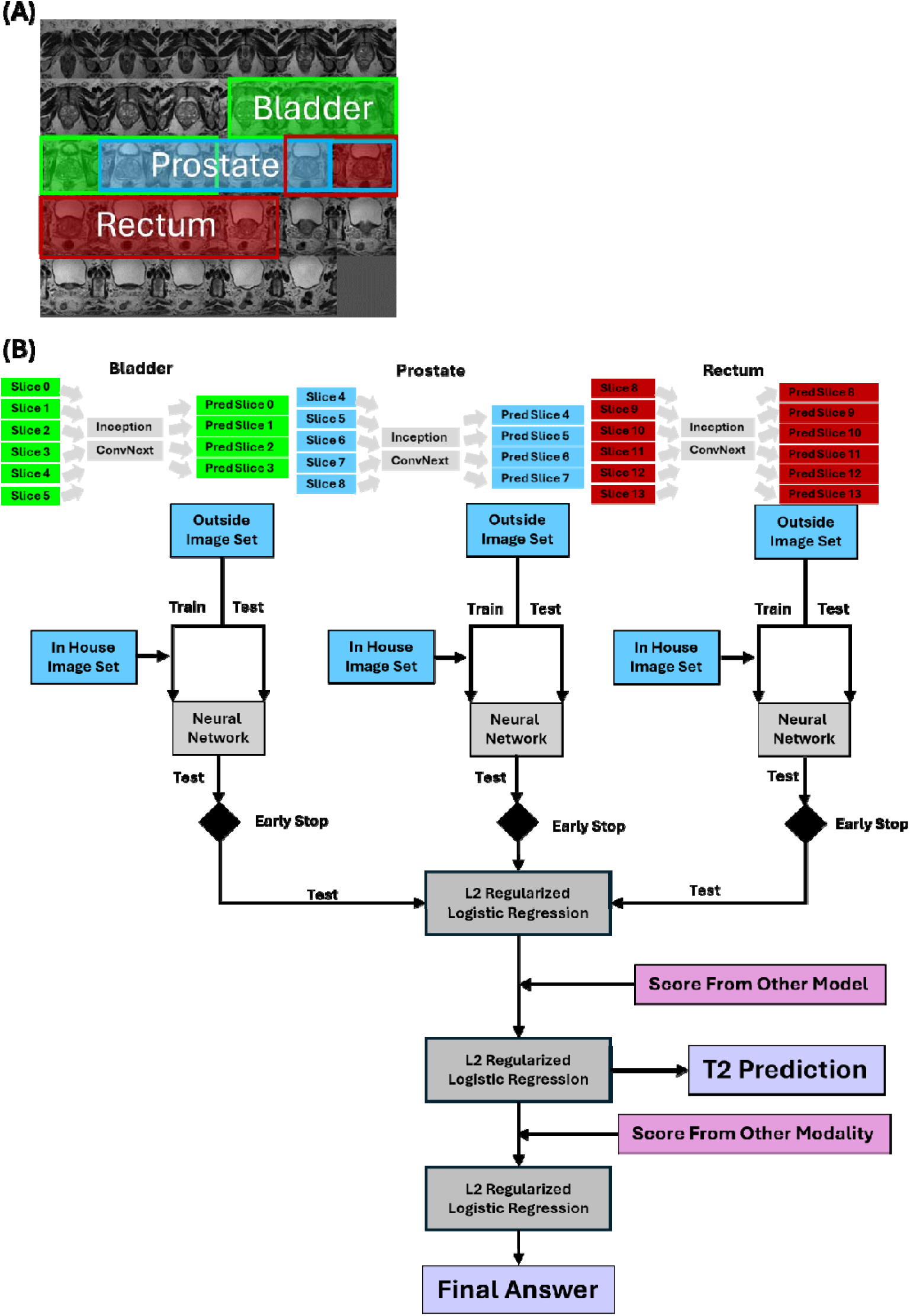
Schematic overview of the multi-stage deep learning approach for prostate MRI quality assessment. (A) Representative T2-weighted axial slice showing the three anatomically relevant regions (bladder, prostate, and rectum) used for analysis. (B) Workflow diagram illustrating the hierarchical integration of neural networks. Each anatomical region is processed by both Inception and ConvNext networks, with predictions combined through L2 regularized logistic regression. The model incorporates scores from other modalities and models to generate T2 predictions and final quality assessments, with early stopping mechanisms at each neural network stage.

For each region and modality (T2 and ADC), we trained two complementary neural networks: Google Inception (15, 16), selected for its multi-scale perception through inception modules, and ConvNext (17), which provides transformer-like performance in a CNN architecture. Both networks were trained using CORN (Consistent Rank Logits) ordinal regression to predict quality scores on a three-point scale (18). The networks were trained with adaptive sharpness-aware minimization (ASAM) (19) using a cosine annealing learning rate schedule (initial lr=5e-5, minimum lr=1e-6) and L2 regularization (weight decay=1e-2) with the Adam optimizer (20). To address class imbalance (21), we implemented a weighted loss function where weights were inversely proportional to class frequency and multiplied the weight of the lowest quality category by 5 to enhance sensitivity for problematic scans. Data augmentation included random horizontal flipping, ±10° rotations, contrast/brightness adjustments, and cutout regularization (22) with a dropout rate of 0.90. Images were preprocessed using CLAHE (Contrast Limited Adaptive Histogram Equalization) (23) with a clip limit of 2.0 and 8x8 tile size, then center-cropped to 164 pixels and resized to 299x299. Early stopping based on maximum AUC in the test set and adaptive sharpness aware minimization (ASAM) was used during training to prevent overfitting (19).

The final model employed a hierarchical regression approach to combine predictions across networks, regions, and modalities. For each of the three anatomical regions, the three-point quality scores were collapsed into a binary classification (non-diagnostic vs diagnostic) before predictions from both Inception and ConvNext networks were combined using L2 regularized logistic regression (liblinear solver, balanced class weights, 5-fold cross-validation for parameter selection). The regional scores were then merged to create modality-specific predictions using another L2 regularized logistic regression model. Finally, T2 and ADC modality scores were combined through regularized regression to produce a single quality assessment score. To ensure unbiased performance assessment, the regression models were trained on the test set data (which was independent from the neural network training set) and evaluated on a separate validation set with no patient overlap as described above.

### Rectal Cross-Sectional Area Analysis

While the neural network achieved robust predictive performance, its ‘black box’ nature makes it challenging to derive explicit quality control procedures from its decisions. We therefore investigated alternative metrics directly linked to specific imaging artifacts. We focused on susceptibility-induced distortion, which is particularly evident near the rectum. Rectal cross-sectional area was measured in every patient using the Universal Segmenter (UniverSeg) model using a soft classification scheme trained on 40 support images manually segmented images. Measurements were performed on the central slice of both T2 and ADC images. The Student t test was used to compare rectal cross-section between quality groups.

### Statistical Analysis

The influence of individual acquisition parameters on image quality was assessed by a multi-pronged statistical approach using the binary classification scheme. The Kruskal-Wallis non-parametric rank test was used to compare median values across different quality categories. The Kolmogorov-Smirnov test assessed the overall distribution of parameters between passing and failing scans. Finally, piecewise logistic regression identified potential thresholds in acquisition parameters that could serve as quality control indicators. Dunn’s multiple comparisons test was used to reduce false positives to reduce false positives from multiple hypothesis testing. P-values were adjusted independently for each statistical test. Statistical calculations were performed using GraphPad Prism 10.1.2(324), except for piecewise regression which used the segmented R package (version 2-12).

## Results

### Quality Analysis of the Study Population

Examples of high- and low-quality MRI scans for T2 images and ADC maps are shown in Figure 1A. There was a significant disparity in image quality between institutions, with 25% of ADC maps and 15% of T2 image sets from outside institutions rated as unsatisfactory, compared to 8% and 3% for the in-house images, respectively (Fig. 1C). To understand these differences, we analyzed representative high- and low-quality scans (Fig. 1A) and categorized the most common artifacts affecting image quality. Our analysis identified three primary types of artifacts in non-diagnostic images. Consistent with previous reports, (6, 24) susceptibility artifacts from rectal gas could be found in the majority of the clinically non-diagnostic ADC images (Fig. S1A-C). While these artifacts were common in outside institution scans, they were rarely observed in in-house images. Low contrast affecting both DWI and ADC maps represented the second most common artifact type (Fig. S1D-F), while aliasing from improper field of view was observed less frequently (Fig. S1G-I). T2W image quality issues were less common overall (15% external vs 3% in-house) and primarily manifested as breathing-related motion artifacts (Fig. S1J).

Notably, high image quality in one modality did not guarantee similar quality in another. The quality of the preceding T2 scan was only moderately correlated with the quality of the ADC map during the same visit (r∼0.4 for both sites) (Fig. 1C). This disconnect between T2 and ADC quality raises the question of whether these imaging failures reflect inherent patient characteristics or technical factors. To address this question, we examined the pattern of successful and failed scans across institutions:

### Impact of Patient Anatomy on Image Quality

The majority of patients were successfully imaged, with only 4 out of 235 T2 images and 9 out of 235 ADC images receiving a non-diagnostic quality score at both sites (Fig. 1B). This low number indicates that it was possible to image most patients successfully at least at one institution. There was a low but non-zero correlation (r∼0.2) between paired scans of the same patient at different sites for both modalities (Fig. 1C), suggesting that while intrinsic patient characteristics played a role, they were not the primary determinant of overall MRI scan quality. However, the observed number of double failures for T2 images exceeded the expected number under the null hypothesis of independence (Fisher’s exact test, p<0.001, ADC p=0.07), suggesting that failures were not entirely independent events. Specifically, a review of representative slices from all cases where imaging was significantly compromised on both in-house and external sites suggested bladder distension, sometimes coincident with or secondary to benign prostatic hyperplasia, was particularly problematic for both T2 and ADC images (Fig. S2). Nevertheless, these cases represented exceptions rather than the norm, indicating that patient anatomy was not the main factor influencing MRI scan quality. This implies that with proper procedures, the vast majority of patients can be successfully scanned.

### Impact of Technical Parameters on Image Quality

As patient anatomy was found not to predominantly influence MRI scan quality, we further interrogated technical acquisition parameters to see if this played key roles. Since the in-house dataset did not have significant image acquisition diversity, we focused our analysis on the multicenter dataset to better understand the role of technical parameters across diverse imaging environments. Our analysis of the multicenter external data confirmed previous findings, no association existed between PI-RADSv2 adherence and MRI quality for both T2 and ADC modalities (Fig. 3C and F). We then conducted a comprehensive statistical analysis of individual acquisition parameters from DICOM files in our multicenter dataset using three complementary approaches: the Kruskal-Wallis test to examine median differences across quality categories on a 3-point scale, the Kolmogorov-Smirnov test to compare parameter distributions between passing and failing scans (Figs 3A and B), and piecewise logistic regression to identify potential quality control thresholds. Despite this comprehensive analysis, no T2 acquisition parameter except a horizontal phase encoding direction in logistic regression emerged as statistically significant (p<0.001, AUC=0. 0.6735), suggesting that T2 image quality is not largely determined by any single technical parameter in isolation.

**Figure 3.**
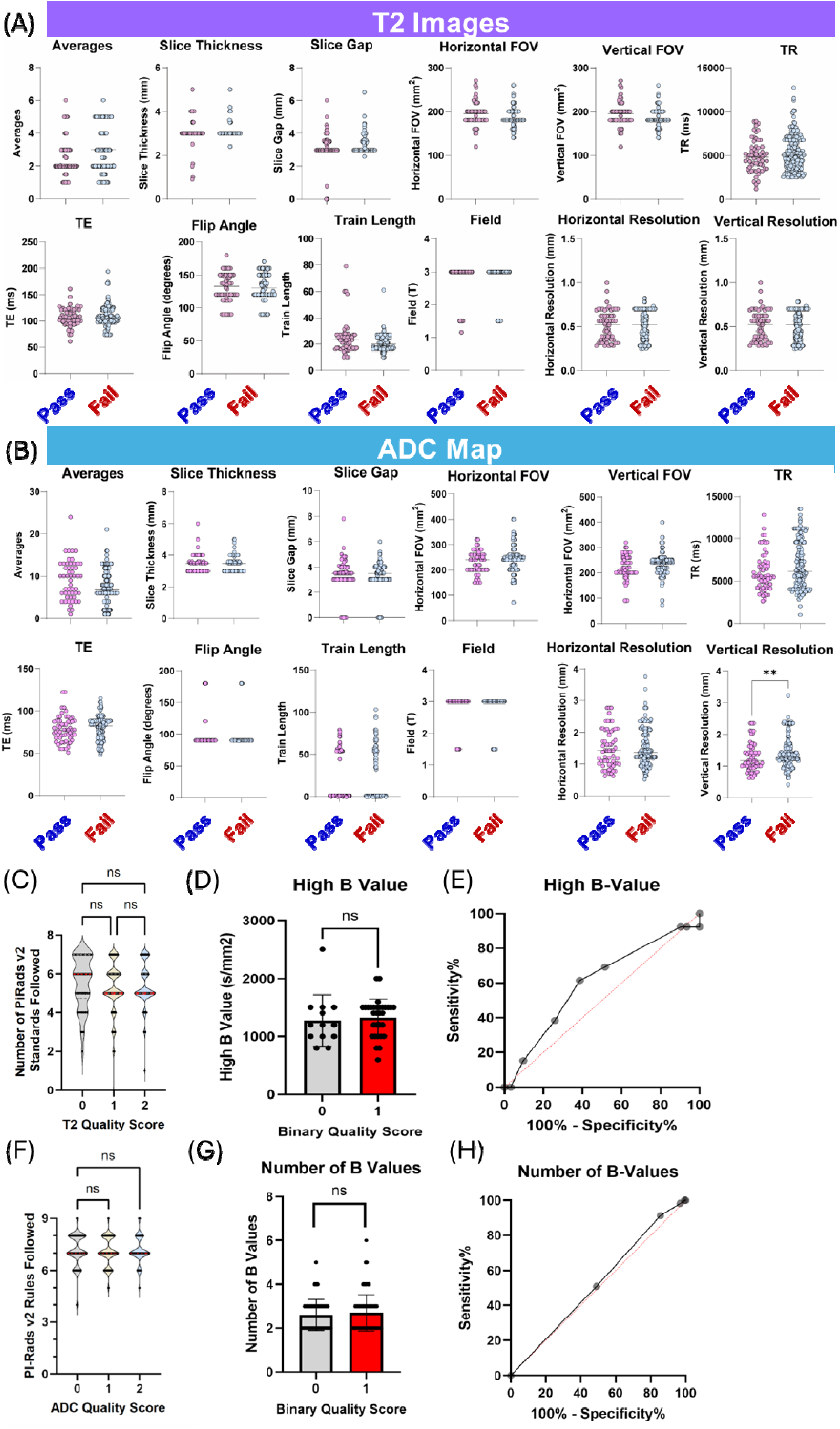
(A and B) Pairwise comparison of acquisition parameters between diagnostic (“Pass”) and non-diagnostic (“Fail”) quality images for T2-weighted images (A) and ADC maps (B). Except for the vertical resolution in ADC maps, no parameter was statistically significant in the multicenter dataset by Kolmogorov-Smirnov test with Holm-Sidak correction for multiple comparisons. (C) Association between PI-RADS v2 standards adherence and reader quality scores for T2-weighted images. (D) Pairwise comparison of the high B value of the ADC map. (E) ROC curve of the high B value (AUC=0.60) (F) Association between PI-RADS v2.1 standards adherence and reader quality scores for T2-weighted images. (G) Pairwise comparison of the number of B values used to make the ADC map. (H) ROC curve of the number of B values (AUC=0.52)

For ADC maps, results were similar, with only the vertical resolution (KS test, p=0.006, AUC= 0.6348) emerging as statistically significant. We specifically did not see a statistically significant difference with factors that had previously been shown to improve MRI quality in direct comparisons including the number of B values (Fig.3D and E),(25) the FOV in either direction (Fig. 3A and B)(26, 27). or the maximum B value (Fig.3G and H),(28–30) although the analysis of the latter was hampered by the low retrieval rate from the DICOM files (19%). This disparity likely stemmed from a combination of technical acquisition differences and patient preparation steps, making it challenging to isolate specific factors without considering potential interactions between imaging parameters.

### Deep Learning-Based Multi-Stage Quality Assessment

Results of our quality assessment neural network (Fig. 2) are shown in Table 1 and Fig. 4. The first section shows the results of the model of each cross-validation fold against the validation set, while the bottom section is the best performing model in the test set against the validation set. In the multicenter validation set, T2 image analysis achieved 65±13% sensitivity and 44±10% positive predictive value (PPV), with the best model reaching 83% sensitivity and 50%, with a notably high negative predictive value (NPV) of 90%. The ADC map assessment showed comparable performance (67% sensitivity, 62% PPV, 88% NPV), while the combined T2/ADC analysis yielded the best overall performance with 83% sensitivity, 50% PPV, and 93% NPV. Importantly, the T2 assessment step, which allows for immediate corrective actions, maintained high sensitivity (83%) and NPV (90%), enables early identification of potential quality issues. The model showed even stronger performance on the in-house dataset, achieving >90% accuracy across all modalities (T2: 92%, ADC: 94%, Combined: 94%), despite being trained exclusively on multicenter data. Additional experiments training ConvNext models on either multicenter or in house data alone showed no difference in accuracy when tested on data from the other site. The model showed similar performance whether trained on multicenter data and tested on in-house data or vice versa, with no significant difference in accuracy compared to within-site testing (training and testing on the same dataset, Table 2). This suggests robust generalization across different imaging environments. This robust generalization, combined with the high NPV across all conditions, suggests the model could effectively screen out poor-quality scans while minimizing unnecessary workflow interruptions.

**Figure 4.**
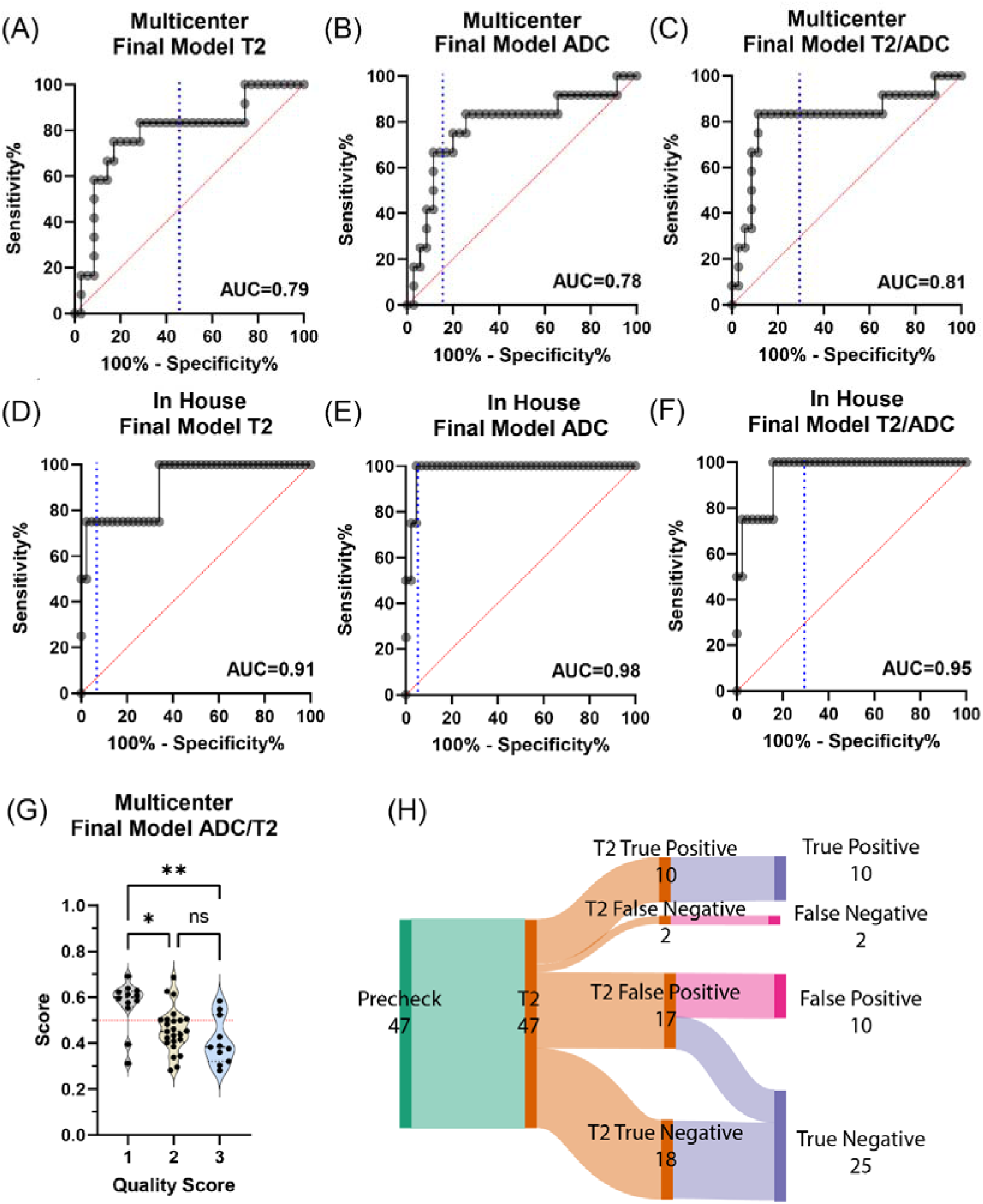
Performance metrics of the quality prediction model based on an independent validation set. **(A-C)** ROC curves for the multicenter dataset showing T2, ADC, and combined T2/ADC model performance respectively. Blue dashed line indicates the cutoff. **(D-F)** Corresponding ROC curves for the in-house dataset demonstrating stronger performance across all modalities. (G) Distribution of model scores across quality categories in the multicenter dataset (*p<0.05, **p<0.01). **(H)** Sankey diagram illustrating the flow of cases through the quality assessment pipeline, showing the distribution of true/false positives and negatives at each stage

**Table 1:** Model performance metrics for quality prediction across different training and testing scenarios. NPV: Negative Predictive Value, PPV: Positive Predictive Value, AUC: Area Under the ROC Curve.

| Multicenter |  |  |  |  |  |  |
| --- | --- | --- | --- | --- | --- | --- |
| From ... | Accuracy | NPV | PPV | Specificity | Sensitivity | AUC |
| T2 | 69 ± 8% | 86 ± 6% | 44 ± 10% | 70 ± 13% | 65 ± 13% | 0.72 ± 0.05 |
| ADC | 69 ± 7% | 88 ± 3% | 45 ± 3% | 69 ± 11% | 72 ± 11% | 0.79 ± 0.02 |
| Combined | 71 ± 6% | 89 ± 3% | 46 ± 7% | 69 ± 8% | 75 ± 6% | 0.78 ± 0.02 |
| Multicenter – Selected Model |  |  |  |  |  |  |
| From ... | Accuracy | NPV | PPV | Specificity | Sensitivity | AUC |
| T2 | 60% | 90% | 37% | 51% | 83% | 0.79 |
| ADC | 81% | 88% | 62% | 86% | 67% | 0.78 |
| Combined | 74% | 93% | 50% | 71% | 83% | 0.81 |
| In House |  |  |  |  |  |  |
| From ... | Accuracy | NPV | PPV | Specificity | Sensitivity | AUC |
| T2 | 90 ± 1% | 98 ± 0% | 46 ± 4% | 92 ± 1% | 75 ± 0% | 0.90 ± 0.02 |
| ADC | 94 ± 3% | 99 ± 1% | 61 ± 14% | 95 ± 3% | 90 ± 14% | 0.99 ± 0.01 |
| Combined | 94 ± 2% | 98 ± 0% | 64 ± 11% | 96 ± 2% | 75 ± 0% | 0.96 ± 0.02 |
| In House – Selected Model |  |  |  |  |  |  |
| From ... | Accuracy | NPV | PPV | Specificity | Sensitivity | AUC |
| T2 | 92% | 98% | 50% | 93% | 75% | 0.91 |
| ADC | 94% | 98% | 60% | 95% | 75% | 0.98 |
| Combined | 94% | 98% | 60% | 95% | 75% | 0.95 |

**Table 2.**
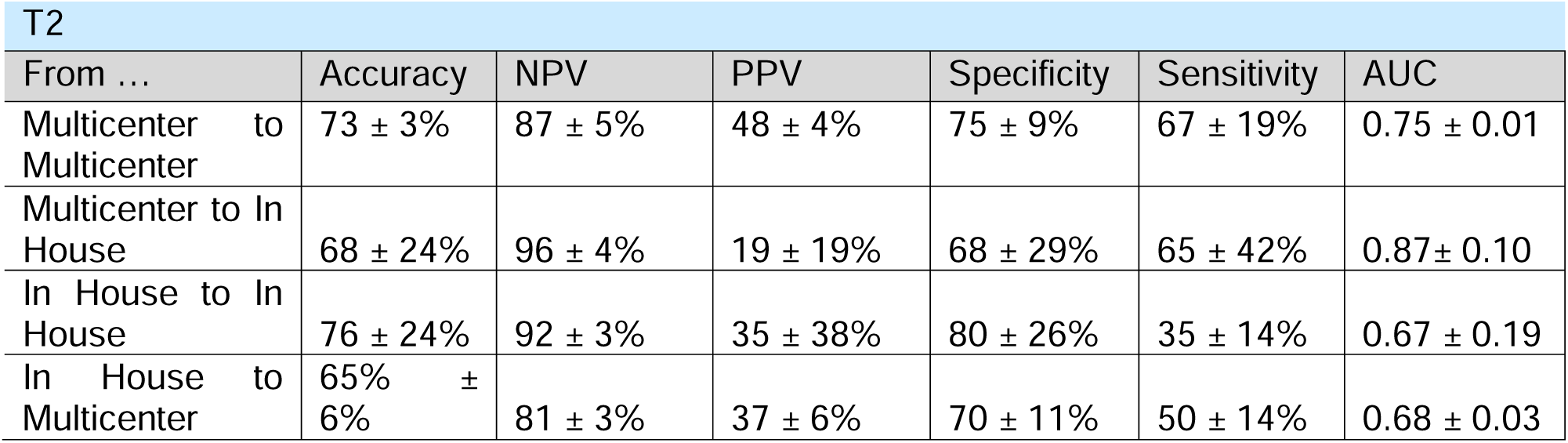
Cross-site validation of T2 image quality prediction. Performance metrics (mean ± standard deviation) for models trained and tested on different combinations of multicenter and in-house datasets. ‘From … to’ indicates training and test sets respectively. NPV: Negative Predictive Value, PPV: Positive Predictive Value, AUC: Area Under the ROC Curve.

| T2 |  |  |  |  |  |  |
| --- | --- | --- | --- | --- | --- | --- |
| From ... | Accuracy | NPV | PPV | Specificity | Sensitivity | AUC |
| Multicenter to Multicenter | 73 ± 3% | 87 ± 5% | 48 ± 4% | 75 ± 9% | 67 ± 19% | 0.75 ± 0.01 |
| Multicenter to In House | 68 ± 24% | 96 ± 4% | 19 ± 19% | 68 ± 29% | 65 ± 42% | 0.87 ± 0.10 |
| In House to In House | 76 ± 24% | 92 ± 3% | 35 ± 38% | 80 ± 26% | 35 ± 14% | 0.67 ± 0.19 |
| In House to Multicenter | 65% ± 6% | 81 ± 3% | 37 ± 6% | 70 ± 11% | 50 ± 14% | 0.68 ± 0.03 |

### Rectal Cross-Sectional Area as a Simple, Interpretable Quality Metric

Using the UniverSeg (31) to automatically measure rectal cross-sectional area in the central slice, we found significant correlations with image quality (Fig. 5) (32). Rectal cross-sectional area on T2 images showed a statistically significant difference between the lowest and highest quality ADC maps (Kruskal-Wallis, p=0.006). This difference was even more pronounced when analyzing ADC maps retrospectively, with significant differences between the lowest and all other quality categories (Kruskal-Wallis, p=0.008 and p<0.0001) (Fig. 5). Logistic regression identified specific breakpoints at 618 mm² for T2 and 760 mm² for ADC maps (AUC 0.65 and 0.69 respectively), suggesting potential threshold values for quality control procedures.

**Figure 5.**
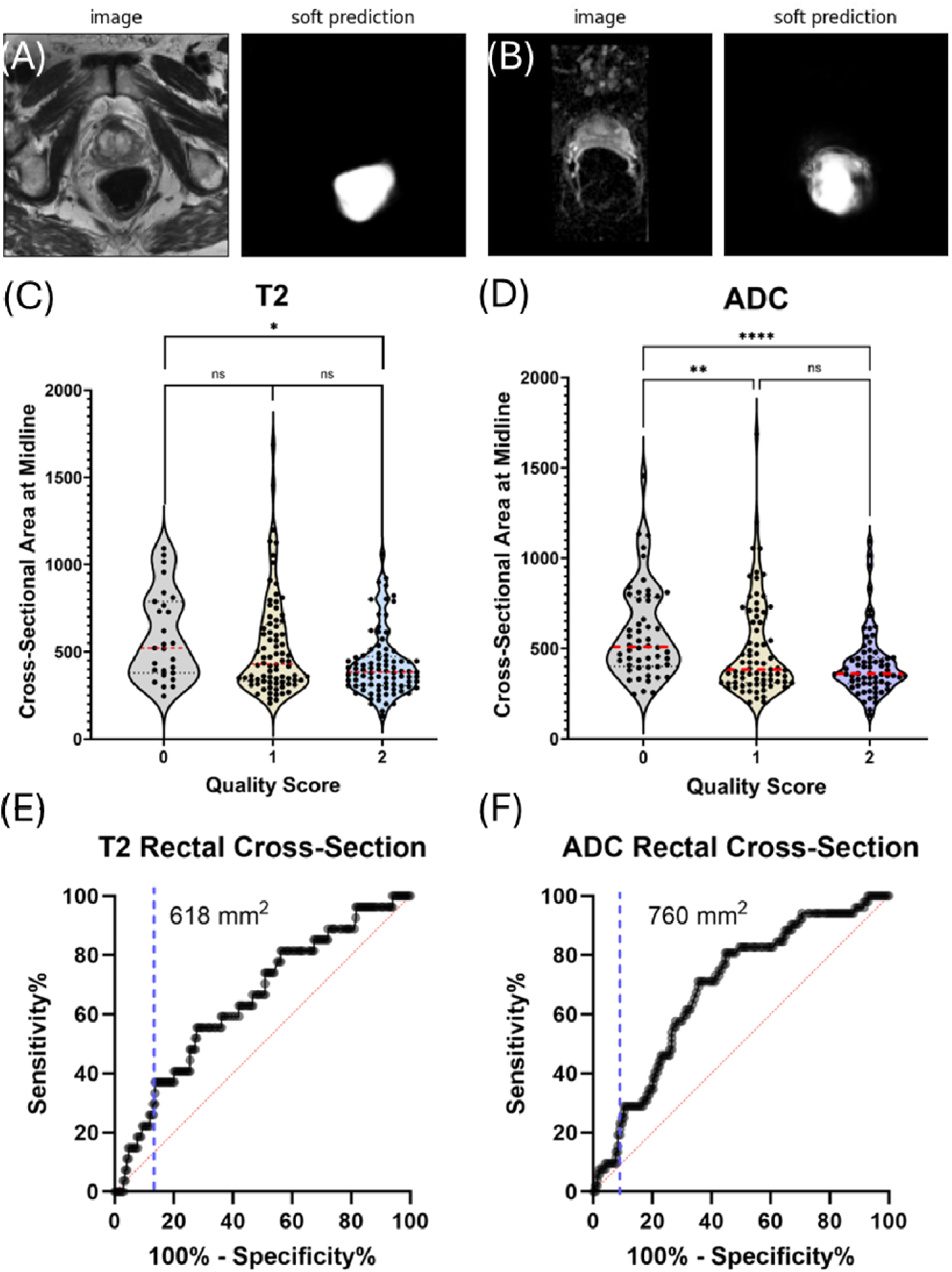
Automated rectal segmentation and its relationship to image quality. **(A)** Example T2 image with corresponding soft prediction from Universal Segmenter. (B) Example ADC map with corresponding segmentation prediction. **(C)** Violin plots showing the distribution of rectal cross-sectional area across T2 image quality scores, with significant difference between lowest and highest quality categories (*p=0.006). **(D)** Similar plots for ADC maps showing significant differences between lowest and other quality categories (**p=0.008, ****p<0.0001). **(E,F)** ROC curves for predicting image quality based on rectal cross-sectional area measurements, with optimal thresholds identified at 618 mm² for T2 and 760 mm² for ADC maps (AUC 0.65 and 0.69 respectively)

## Discussion

The performance of deep learning models trained on data from a single clinical site often deteriorates when deployed in a multicenter environment. (33–36) Our multi-center neural network showed performance comparable to single-site models for ADC quality prediction (10, 32, 37), though direct comparisons are challenging due to substantial inter-study variations in poor-quality image rates and validation procedures. Our in-house accuracy approximates the performance of the best single site model, providing strong validation for our approach.(10) The top-performing model in the literature is the single-site model by Cipollari et al., which achieved 83% sensitivity and 100% specificity in a similar non-diagnostic vs. diagnostic binary classification system.(10) This is comparable to our accuracy when tested on in-house data (75% sensitivity and 96% specificity) but exceeds our multi-center accuracy (83% sensitivity and 69% specificity) (Table 1). The lower accuracy in our multi-center data analysis may be attributed to the increased variability in imaging protocols across different institutions.

This variability could limit the model’s ability to learn protocol-specific artifacts as relevant features, leading to a potential trade-off between site-specific accuracy and generalizability (34, 38). Consistent with this hypothesis, Alis et al.’s multi-center study, the only other study of its kind, showed a lower overall accuracy compared to single-center studies when translated to a binary system. This suggests that increased protocol variability may negatively impact the model’s overall performance. Overall, our model maintained robust accuracy across multiple centers, even in the face of significant protocol variability across institutions.

While aligning with previous studies in terms of overall performance, our model extends automatic quality assessment’s potential by predicting ADC quality directly from T2 images, an approach not yet widely explored (32). Predicting ADC quality before ADC acquisition offers a crucial advantage. T2 images are routinely acquired first, enabling an immediate assessment of potential quality issues before proceeding with the potentially longer ADC scan. This allows for timely interventions like protocol adjustments or patient repositioning, optimizing workflow efficiency and potentially reducing unnecessary ADC acquisitions (Fig 6). To take advantage of this early quality assessment capability, an efficient system requires both high sensitivity at the T2 stage to identify potential issues and a high negative predictive value, given that most scans are of acceptable quality. Our model demonstrated robust performance across different imaging environments (Fig. 4 and Table 1), achieving 83% sensitivity with 90% NPV in the multicenter validation set, while showing exceptional discriminative power on in-house data (AUC>0.90 for all modalities). This combination of strong cross-site generalization and high negative predictive value suggests the model could effectively screen out poor-quality scans while minimizing unnecessary workflow interruptions.

**Figure 6:**
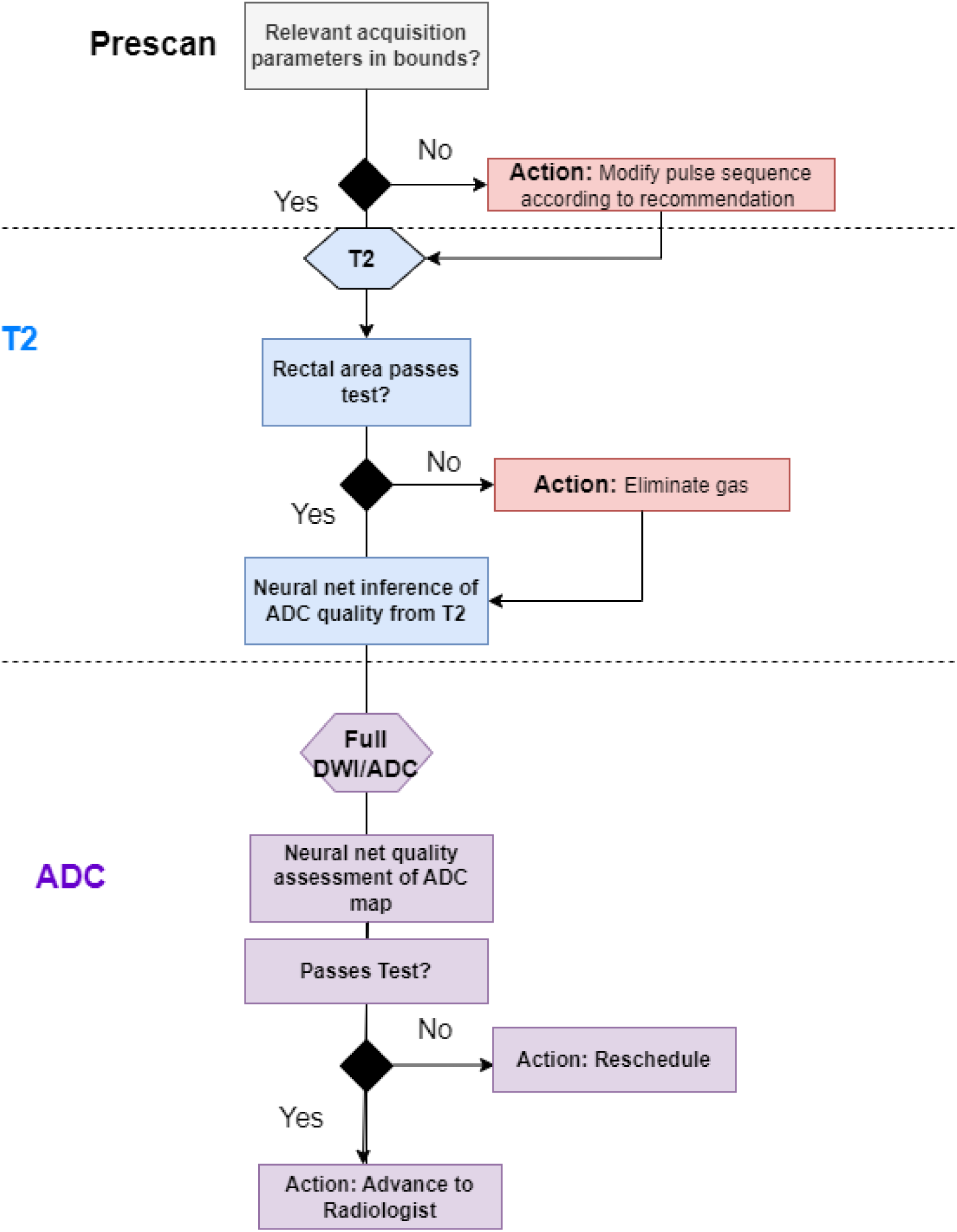
Proposed decision workflow for prostate MRI quality control. The process begins with prescan parameter verification, followed by T2-weighted image acquisition and quality assessment. Neural network inference predicts potential ADC quality from T2 images before proceeding to full DWI/ADC acquisition. Quality control decision points (diamonds) trigger specific interventions (red boxes) when quality thresholds are not met, with final advancement to radiologist review only after passing all quality checks.

Beyond the immediate benefits of T2-based prediction for optimizing workflow and resource allocation, a deeper understanding of factors influencing T2 and ADC quality can further refine our approach and identify specific targets for intervention. One prominent factor often associated with image quality is signal-to-noise ratio (SNR). Existing research shows mixed results on its true impact on both T2 and ADC images, raising questions about its effectiveness as a major indicator of quality. Belue *et al* found reconstruction by a generative adversarial network strongly increased SNR in T2 images but this increase in SNR rarely led to an increase in perceptual image quality when the reader was blinded to the reconstruction (39). Similarly, Lee et al found no difference in perceptual image quality for T2, DWI, or ADC images after deep learning reconstruction, although high reader variability complicated the statistical interpretation of these results (40). “Noise” as a subjective factor, however, ranks highly in subjective perceptions of MRI image quality (6). This suggests the subjective perception of “noise” may result from other artifacts contributing to low contrast with different physical sources. Low contrast in the presence of high SNR (Fig. S1F) may be the result of the progressive misalignment of voxels due to geometric distortions at higher B values (41), potentially obscuring diagnostic features more critically than traditional noise measurements. This unexpected finding highlights the complex nature of factors influencing perceived image quality and a deeper understanding of image features impacting perceived quality becomes crucial. In general, image quality for both T2 images and ADC maps appears to be largely a transient phenomenon and not heavily dependent on the inherent characteristic of the patient or the technical aspects of the imaging sequence (Fig. 3). In line with this observation, distortion from susceptibility differences from rectal gas, which are especially noticeable near the rectum (Fig. S1D-F), are a significant factor. Interestingly, a simple measure of rectal area in the T2 image correlated relatively well with ADC image quality in the independent validation set (Fig. 5C and E), potentially offering another readily available indicator that can be used as either an alternative indicator or as a readily available marker that could guide interventions (32).

Some limitations of our study should be acknowledged. While our model demonstrates promising results in predicting ADC quality directly from T2 images, it’s crucial to acknowledge limitations in fully understanding the specific features it relies on for its predictions. Unlike established metrics like SNR, which have a clear physical basis, our deep learning model operates through complex internal processes, making it challenging to pinpoint the exact image characteristics driving its quality assessment. Second, our model rates the ADC map and not the underlying DWI images, which may be more critical for diagnosis (3). Our quality interpretation was performed by only a single radiologist and interobserver variation analysis is therefore not possible. Finally, our model is based on perceptual quality and not improvement in accuracy in downstream tasks, which may have different requirements. Despite these limitations, our study demonstrates the strong potential of direct T2-based prediction for optimizing prostate MRI workflow and future research can address these limitations to further refine this approach (32).

In conclusion, we developed an interpretable AI model, which jointly utilizes T2 images and ADC maps to predict non-diagnostic ADC maps. Although our model demonstrated reasonable performance in a multi-center dataset, further evaluation across diverse clinical settings is needed to confirm its potential benefit in practice (42).

## Data Availability Statement

The code used in this study is publicly available at https://github.com/jbrender/QCMRI. The imaging datasets used during the current study are available from the corresponding author upon reasonable request.

---

**Figure S1:**
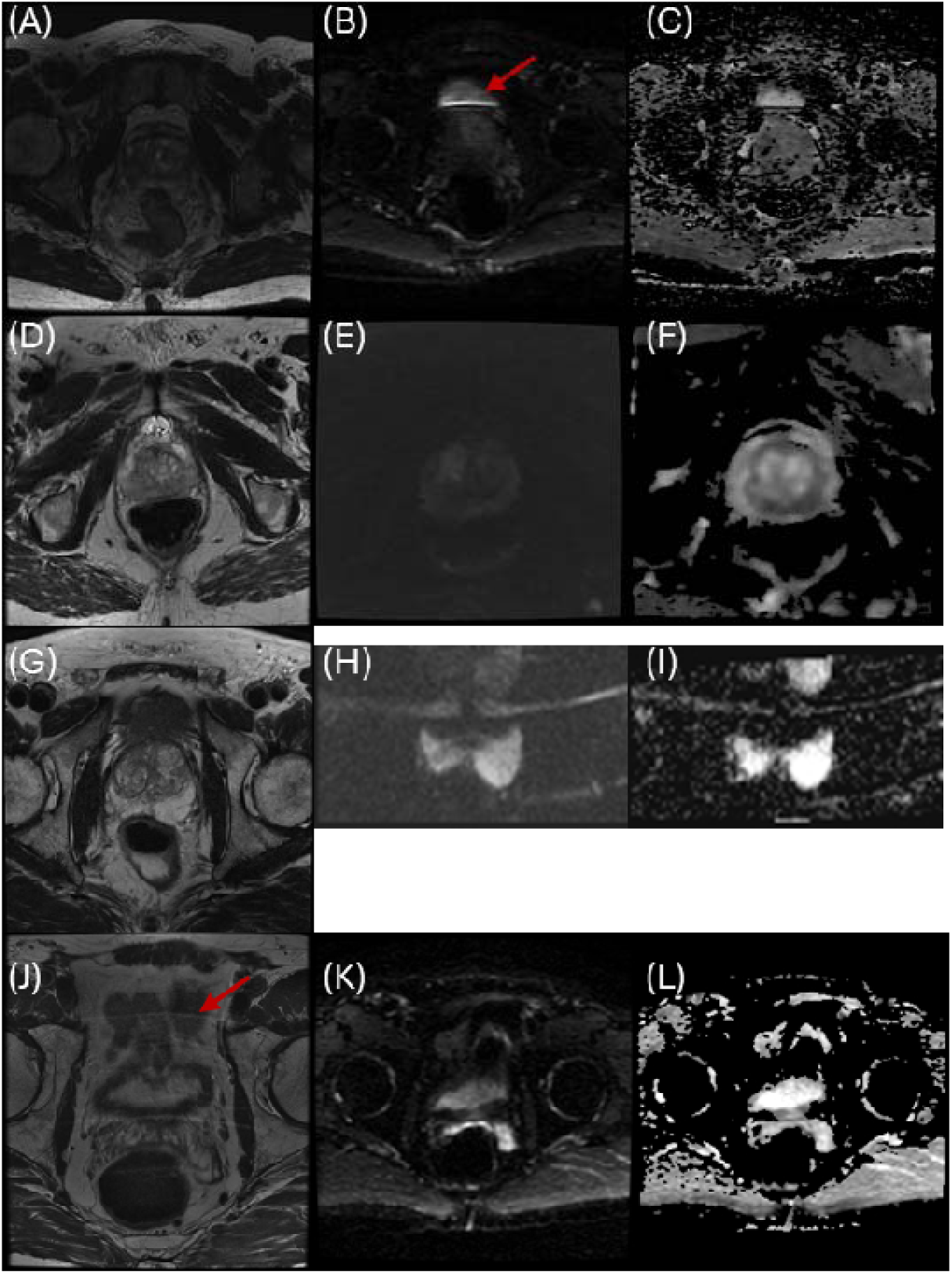
Representative examples of common image quality issues in prostate MRI. **(A-C)** Susceptibility artifacts due to rectal gas: T2-weighted image showing subtle geometric distortion **(A)**, corresponding high b-value DWI with signal pileup at the prostate-rectum interface **(B, arrow)**, and ADC map demonstrating signal dropout and reduced contrast **(C)**. **(D-F)** Low contrast artifacts: T2-weighted image with normal anatomy **(D)**, DWI showing poor zone differentiation **(E)**, and corresponding low-contrast ADC map **(F)**. **(G-I)** Aliasing artifacts from reduced field of view: Normal T2-weighted image with expanded FOV **(G)**, reduced FOV DWI with bright band artifacts **(H)**, and ADC map showing wraparound effects **(I)**. **(J-L)** Motion artifact apparent in the T2 image **(J, arrow)**

**Figure S2:**
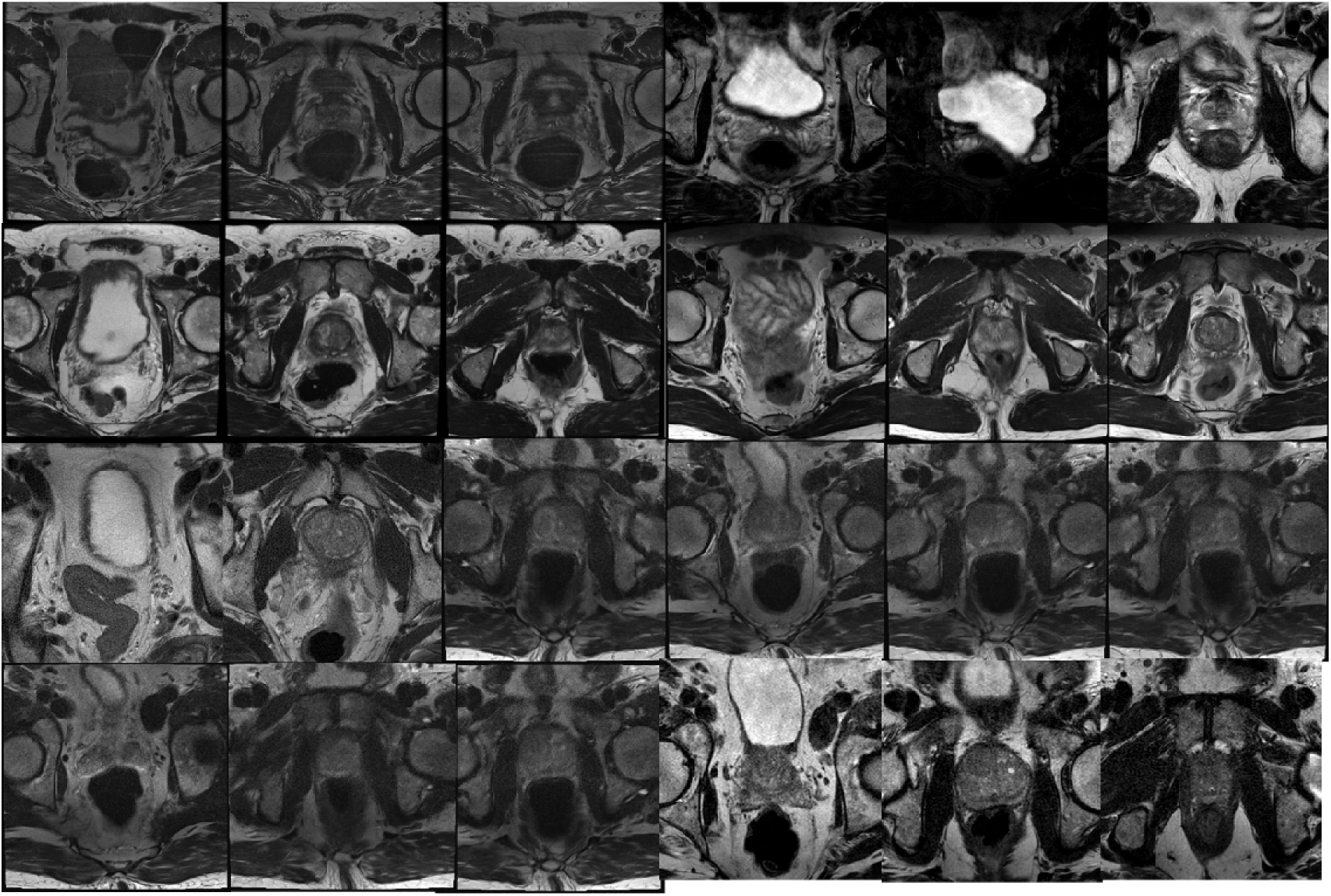
Representative T2-weighted images from cases with non-diagnostic quality at both in-house (left 3 images) and external sites (right). Each row shows consecutive axial slices through the prostate demonstrating how bladder distension, often associated with benign prostatic hyperplasia, compromises image quality. Note the distorted anatomy and signal inhomogeneity across multiple slices, making these studies clinically uninterpretable

**Figure S3:**
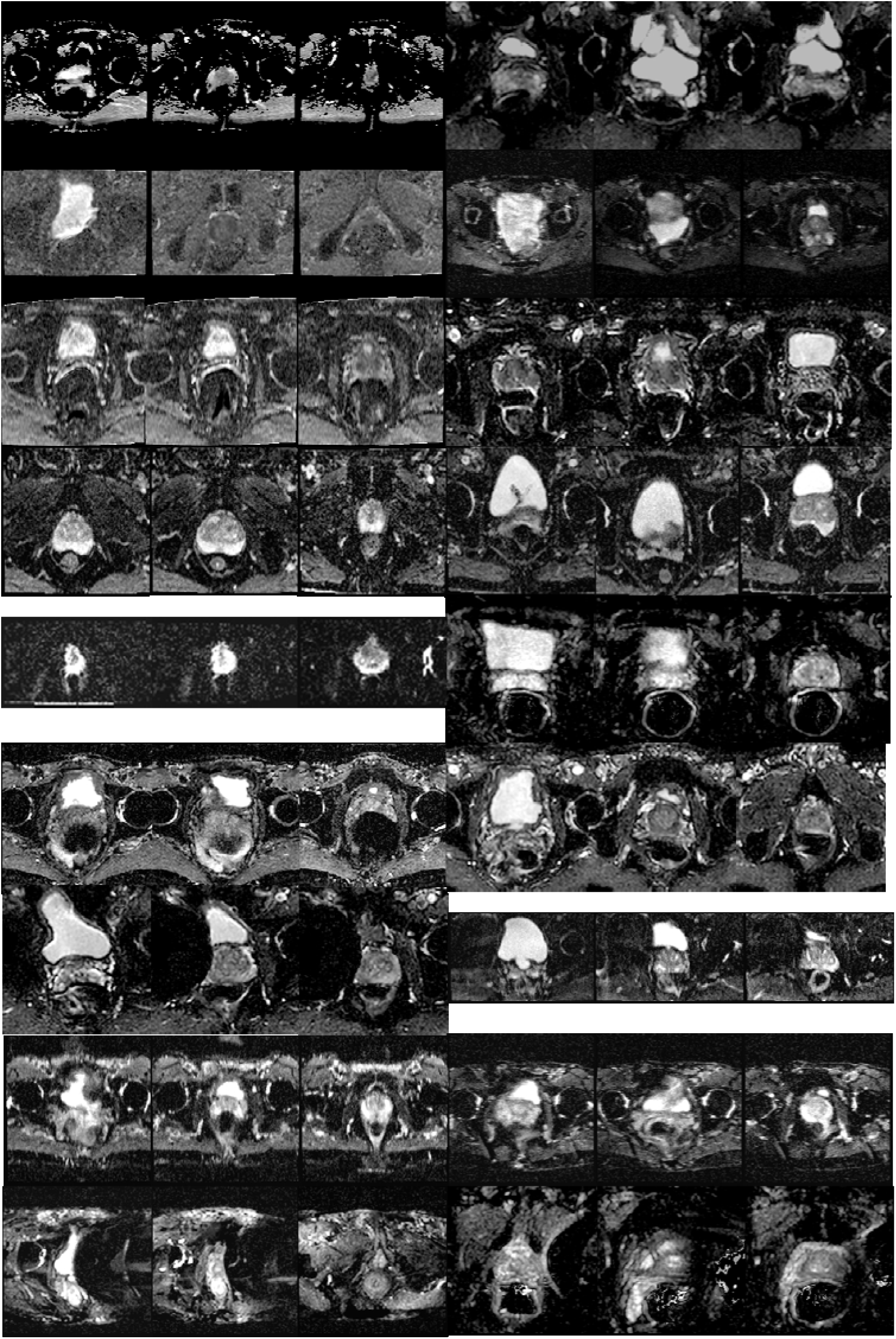
Representative ADC maps from cases with non-diagnostic quality at both in-house and external sites. Each row shows consecutive axial slices through the prostate, illustrating how bladder distension, often in conjunction with benign prostatic hyperplasia, leads to significant image degradation. Note the susceptibility artifacts and geometric distortions across multiple slices that render these studies non-diagnostic.

## Notes

### Competing Interest Statement

The authors have declared no competing interest.

### Clinical Protocols

https://www.clinicaltrials.gov/study/NCT03354416

### Funding Statement

This work was supported by the intramural research programs of the Center for Cancer Research, NCI

### Author Declarations

National Cancer Institute Central Institutional Review Board of the National Cancer Institute gave ethical approval for this work

## References

1. Barrett T, de Rooij M, Giganti F, Allen C, Barentsz JO, Padhani AR. Quality checkpoints in the MRI-directed prostate cancer diagnostic pathway. Nat Rev Urol 2023;20(1):9–22. doi: 10.1038/s41585-022-00648-4

2. Windisch O, Benamran D, Dariane C, Favre MM, Djouhri M, Chevalier M, Guillaume B, Oderda M, Gatti M, Faletti R, Colinet V, Lefebvre Y, Bodard S, Diamand R, Fiard G. Role of the Prostate Imaging Quality PI-QUAL Score for Prostate Magnetic Resonance Image Quality in Pathological Upstaging After Radical Prostatectomy: A Multicentre European Study. Eur Urol Open Sci 2023;47:94–101. doi: 10.1016/j.euros.2022.11.013

3. Weinreb JC, Barentsz JO, Choyke PL, Cornud F, Haider MA, Macura KJ, Margolis D, Schnall MD, Shtern F, Tempany CM, Thoeny HC, Verma S. PI-RADS Prostate Imaging - Reporting and Data System: 2015, Version 2. Eur Urol 2016;69(1):16–40. doi: 10.1016/j.eururo.2015.08.052

4. Giganti F, Allen C, Emberton M, Moore CM, Kasivisvanathan V, group Ps. Prostate Imaging Quality (PI-QUAL): A New Quality Control Scoring System for Multiparametric Magnetic Resonance Imaging of the Prostate from the PRECISION trial. Eur Urol Oncol 2020;3(5):615–619. doi: 10.1016/j.euo.2020.06.007

5. Giganti F, Ng A, Asif A, Chan VW, Rossiter M, Nathan A, Khetrapal P, Dickinson L, Punwani S, Brew-Graves C, Freeman A, Emberton M, Moore CM, Allen C, Kasivisvanathan V, Group PQI. Global Variation in Magnetic Resonance Imaging Quality of the Prostate. Radiology 2023;309(1):e231130. doi: 10.1148/radiol.231130

6. Sackett J, Shih JH, Reese SE, Brender JR, Harmon SA, Barrett T, Coskun M, Madariaga M, Marko J, Law YM, Turkbey EB, Mehralivand S, Sanford T, Lay N, Pinto PA, Wood BJ, Choyke PL, Turkbey B. Quality of Prostate MRI: Is the PI-RADS Standard Sufficient? Acad Radiol 2021;28(2):199–207. doi: 10.1016/j.acra.2020.01.031

7. Sadri AR, Janowczyk A, Zhou R, Verma R, Beig N, Antunes J, Madabhushi A, Tiwari P, Viswanath SE. Technical Note: MRQy - An open-source tool for quality control of MR imaging data. Med Phys 2020;47(12):6029–6038. doi: 10.1002/mp.14593

8. Giganti F, Lindner S, Piper JW, Kasivisvanathan V, Emberton M, Moore CM, Allen C. Multiparametric prostate MRI quality assessment using a semi-automated PI-QUAL software program. Eur Radiol Exp 2021;5(1):48. doi: 10.1186/s41747-021-00245-x

9. Lin Y, Yilmaz EC, Belue MJ, Turkbey B. Prostate MRI and image Quality: It is time to take stock. Eur J Radiol 2023;161:110757. doi: 10.1016/j.ejrad.2023.110757

10. Cipollari S, Guarrasi V, Pecoraro M, Bicchetti M, Messina E, Farina L, Paci P, Catalano C, Panebianco V. Convolutional Neural Networks for Automated Classification of Prostate Multiparametric Magnetic Resonance Imaging Based on Image Quality. J Magn Reson Imaging 2022;55(2):480–490. doi: 10.1002/jmri.27879

11. Thijssen LCP, de Rooij M, Barentsz JO, Huisman HJ. Radiomics based automated quality assessment for T2W prostate MR images. Eur J Radiol 2023;165:110928. doi: 10.1016/j.ejrad.2023.110928

12. Alis D, Kartal MS, Seker ME, Guroz B, Basar Y, Arslan A, Sirolu S, Kurtcan S, Denizoglu N, Tuzun U, Yildirim D, Oksuz I, Karaarslan E. Deep learning for assessing image quality in bi-parametric prostate MRI: A feasibility study. Eur J Radiol 2023;165:110924. doi: 10.1016/j.ejrad.2023.110924

13. Lin Y, Belue MJ, Yilmaz EC, Harmon SA, An J, Law YM, Hazen L, Garcia C, Merriman KM, Phelps TE, Lay NS, Toubaji A, Merino MJ, Wood BJ, Gurram S, Choyke PL, Pinto PA, Turkbey B. Deep Learning-Based T2-Weighted MR Image Quality Assessment and Its Impact on Prostate Cancer Detection Rates. J Magn Reson Imaging 2024;59(6):2215–2223. doi: 10.1002/jmri.29031

14. de Rooij M, Allen C, Twilt JJ, Thijssen LCP, Asbach P, Barrett T, Brembilla G, Emberton M, Gupta RT, Haider MA, Kasivisvanathan V, Logager V, Moore CM, Padhani AR, Panebianco V, Puech P, Purysko AS, Renard-Penna R, Richenberg J, Salomon G, Sanguedolce F, Schoots IG, Thony HC, Turkbey B, Villeirs G, Walz J, Barentsz J, Giganti F. PI-QUAL version 2: an update of a standardised scoring system for the assessment of image quality of prostate MRI. Eur Radiol 2024;34(11):7068–7079. doi: 10.1007/s00330-024-10795-4

15. Szegedy C, Vanhoucke V, Ioffe S, Shlens J, Wojna Z. Rethinking the Inception Architecture for Computer Vision. 2016 IEEE Conference on Computer Vision and Pattern Recognition (CVPR) 2016; p. 2818–2826.

16. Szegedy C, Wei L, Yangqing J, Sermanet P, Reed S, Anguelov D, Erhan D, Vanhoucke V, Rabinovich A. Going deeper with convolutions. 2015 IEEE Conference on Computer Vision and Pattern Recognition (CVPR) 2015; p. 1–9.

17. Liu Z, Mao H, Wu CY, Feichtenhofer C, Darrell T, Xie S. A ConvNet for the 2020s. 2022 IEEE/CVF Conference on Computer Vision and Pattern Recognition (CVPR) 2022; p. 11966–11976.

18. Shi X, Cao W, Raschka S. Deep neural networks for rank-consistent ordinal regression based on conditional probabilities. Pattern Analysis and Applications 2023;26(3):941–955. doi: 10.1007/s10044-023-01181-9

19. Kwon J, Kim J, Park H, Choi IK. ASAM: Adaptive Sharpness-Aware Minimization for Scale-Invariant Learning of Deep Neural Networks. In: Marina M, Tong Z, eds. Proceedings of the 38th International Conference on Machine Learning. Proceedings of Machine Learning Research: PMLR, 2021; p. 5905–5914.

20. Kingma DP, Ba J. Adam: A method for stochastic optimization. arXiv preprint arXiv:14126980 2014.

21. Shwartz-Ziv R, Goldblum M, Li YL, Bruss CB, Wilson AG. Simplifying Neural Network Training Under Class Imbalance. arXiv preprint arXiv:231202517 2023.

22. Devries T, Taylor GW. Improved Regularization of Convolutional Neural Networks with Cutout. ArXiv 2017;abs/1708.04552.

23. Pizer SM, Amburn EP, Austin JD, Cromartie R, Geselowitz A, Greer T, ter Haar Romeny B, Zimmerman JB, Zuiderveld K. Adaptive histogram equalization and its variations. Computer Vision, Graphics, and Image Processing 1987;39(3):355–368. doi: 10.1016/S0734-189X(87)80186-X

24. Purysko AS, Zacharias-Andrews K, Tomkins KG, Turkbey IB, Giganti F, Bhargavan-Chatfield M, Larson DB, Collaborative ACRPMIQI. Improving Prostate MR Image Quality in Practice-Initial Results From the ACR Prostate MR Image Quality Improvement Collaborative. J Am Coll Radiol 2024;21(9):1464–1474. doi: 10.1016/j.jacr.2024.04.008

25. Saad LS, de Queiroz Rosas G, de Farias e Melo HJ, Gabriele HAA, Szejnfeld J. ADC mapping with 12 <em>b</em> values: an improved technique for image quality in diffusion prostate MRI. bioRxiv 2019:744961. doi: 10.1101/744961

26. Lawrence EM, Zhang Y, Starekova J, Wang Z, Pirasteh A, Wells SA, Hernando D. Reduced field-of-view and multi-shot DWI acquisition techniques: Prospective evaluation of image quality and distortion reduction in prostate cancer imaging. Magn Reson Imaging 2022;93:108–114. doi: 10.1016/j.mri.2022.08.008

27. Thierfelder KM, Scherr MK, Notohamiprodjo M, Weiss J, Dietrich O, Mueller-Lisse UG, Pfeuffer J, Nikolaou K, Theisen D. Diffusion-weighted MRI of the prostate: advantages of Zoomed EPI with parallel-transmit-accelerated 2D-selective excitation imaging. Eur Radiol 2014;24(12):3233–3241. doi: 10.1007/s00330-014-3347-y

28. Manenti G, Nezzo M, Chegai F, Vasili E, Bonanno E, Simonetti G. DWI of Prostate Cancer: Optimal b-Value in Clinical Practice. Prostate Cancer 2014;2014:868269. doi: 10.1155/2014/868269

29. Li C, Li N, Li Z, Shen L. Diagnostic accuracy of high b-value diffusion weighted imaging for patients with prostate cancer: a diagnostic comprehensive analysis. Aging (Albany NY) 2021;13(12):16404–16424. doi: 10.18632/aging.203164

30. Kim CK, Park BK, Kim B. High-b-value diffusion-weighted imaging at 3 T to detect prostate cancer: comparisons between b values of 1,000 and 2,000 s/mm2. AJR Am J Roentgenol 2010;194(1):W33–37. doi: 10.2214/AJR.09.3004

31. Butoi VI, Ortiz JJG, Ma T, Sabuncu MR, Guttag J, Dalca AV. UniverSeg: Universal Medical Image Segmentation. 2023 IEEE/CVF International Conference on Computer Vision (ICCV): IEEE Computer Society, 2023; p. 21381–21394.

32. Al-Hayali A, Komeili A, Azad A, Sathiadoss P, Schieda N, Ukwatta E. Machine learning based prediction of image quality in prostate MRI using rapid localizer images. J Med Imaging (Bellingham) 2024;11(2):026001. doi: 10.1117/1.JMI.11.2.026001

33. Jarkman S, Karlberg M, Poceviciute M, Boden A, Bandi P, Litjens G, Lundstrom C, Treanor D, van der Laak J. Generalization of Deep Learning in Digital Pathology: Experience in Breast Cancer Metastasis Detection. Cancers (Basel) 2022;14(21). doi: 10.3390/cancers14215424

34. Souza R, Winder A, Stanley EAM, Vigneshwaran V, Camacho M, Camicioli R, Monchi O, Wilms M, Forkert ND. Identifying Biases in a Multicenter MRI Database for Parkinson’s Disease Classification: Is the Disease Classifier a Secret Site Classifier? IEEE J Biomed Health Inform 2024;28(4):2047–2054. doi: 10.1109/JBHI.2024.3352513

35. Sarma KV, Harmon S, Sanford T, Roth HR, Xu Z, Tetreault J, Xu D, Flores MG, Raman AG, Kulkarni R, Wood BJ, Choyke PL, Priester AM, Marks LS, Raman SS, Enzmann D, Turkbey B, Speier W, Arnold CW. Federated learning improves site performance in multicenter deep learning without data sharing. J Am Med Inform Assoc 2021;28(6):1259–1264. doi: 10.1093/jamia/ocaa341

36. Zech JR, Badgeley MA, Liu M, Costa AB, Titano JJ, Oermann EK. Variable generalization performance of a deep learning model to detect pneumonia in chest radiographs: A cross-sectional study. PLoS Med 2018;15(11):e1002683. doi: 10.1371/journal.pmed.1002683

37. Saeed SU, Yan W, Fu Y, Giganti F, Yang Q, Baum ZMC, Rusu M, Fan RE, Sonn GA, Emberton M, Barratt DC, Hu Y. Image quality assessment by overlapping task-specific and task-agnostic measures: application to prostate multiparametric MR images for cancer segmentation. 2022;arXiv:2202.09798. doi: 10.48550/arXiv.2202.09798. Accessed February 01, 2022.

38. Muglia VF, Westphalen AC. Editorial on “Convolutional Neural Networks for Automated Classification of Prostate Multiparametric Magnetic Resonance Imaging Based on Image Quality”. J Magn Reson Imaging 2022;55(2):491–492. doi: 10.1002/jmri.27913

39. Belue MJ, Harmon SA, Masoudi S, Barrett T, Law YM, Purysko AS, Panebianco V, Yilmaz EC, Lin Y, Jadda PK, Raavi S, Wood BJ, Pinto PA, Choyke PL, Turkbey B. Quality of T2-weighted MRI re-acquisition versus deep learning GAN image reconstruction: A multi-reader study. Eur J Radiol 2024;170:111259. doi: 10.1016/j.ejrad.2023.111259

40. Lee KL, Kessler DA, Dezonie S, Chishaya W, Shepherd C, Carmo B, Graves MJ, Barrett T. Assessment of deep learning-based reconstruction on T2-weighted and diffusion-weighted prostate MRI image quality. Eur J Radiol 2023;166:111017. doi: 10.1016/j.ejrad.2023.111017

41. Blackledge MD, Tunariu N, Zungi F, Holbrey R, Orton MR, Ribeiro A, Hughes JC, Scurr ED, Collins DJ, Leach MO, Koh DM. Noise-Corrected, Exponentially Weighted, Diffusion-Weighted MRI (niceDWI) Improves Image Signal Uniformity in Whole-Body Imaging of Metastatic Prostate Cancer. Front Oncol 2020;10:704. doi: 10.3389/fonc.2020.00704

42. Lekadir K, Feragen A, Fofanah AJ, Frangi AF, Buyx A, Emelie A, Lara A, Porras AR, Chan A-W, Navarro A, Glocker B, Botwe BO, Khanal B, Beger B, Wu CC, Cintas C, Langlotz CP, Rueckert D, Mzurikwao D, Fotiadis DI, Zhussupov D, Ferrante E, Meijering E, Weicken E, González FA, Asselbergs FW, Prior F, Krestin GP, Collins G, Tegenaw GS, Kaissis G, Misuraca G, Tsakou G, Dwivedi G, Kondylakis H, Jayakody H, Woodruf HC, Mayer HJ, JWL Aerts H, Walsh I, Chouvarda I, Buvat I, Tributsch I, Rekik I, Duncan J, Kalpathy-Cramer J, Zahir J, Park J, Mongan J, Gichoya JW, Schnabel JA, Kushibar K, Riklund K, Mori K, Marias K, Amugongo LM, Fromont LA, Maier-Hein L, Cerdá Alberich L, Rittner L, Phiri L, Marrakchi-Kacem L, Donoso-Bach L, Martí-Bonmatí L, Cardoso MJ, Bobowicz M, Shabani M, Tsiknakis M, Zuluaga MA, Bielikova M, Fritzsche M-C, Camacho M, Linguraru MG, Wenzel M, De Bruijne M, Tolsgaard MG, Ghassemi M, Ashrafuzzaman M, Goisauf M, Yaqub M, Cano Abadía M, E Mahmoud MM, Elattar M, Rieke N, Papanikolaou N, Lazrak N, Díaz O, Salvado O, Pujol O, Sall O, Guevara P, Gordebeke P, Lambin P, Brown P, Abolmaesumi P, Dou Q, Lu Q, Osuala R, Nakasi R, Zhou SK, Napel S, Colantonio S, Albarqouni S, Joshi S, Carter S, Klein S, E Petersen S, Aussó S, Awate S, Riklin Raviv T, Cook T, Mutsvangwa TEM, Rogers WA, Niessen WJ, Puig-Bosch X, Zeng Y, Mohammed YG, Aquino YSJ, Salahuddin Z, Starmans MPA. FUTURE-AI: International consensus guideline for trustworthy and deployable artificial intelligence in healthcare. 2023;arXiv:2309.12325. doi: 10.48550/arXiv.2309.12325. Accessed August 01, 2023.

